# Estimating Vaccine-Preventable COVID-19 Deaths Under Counterfactual Vaccination Scenarios in the United States

**DOI:** 10.1101/2022.05.19.22275310

**Authors:** Ming Zhong, Meghana Kshirsagar, Richard Johnston, Rahul Dodhia, Tammy Glazer, Allen Kim, Divya Michael, Sameer Nair-Desai, Thomas C. Tsai, Stefanie Friedhoff, Juan M. Lavista Ferres

**Affiliations:** Microsoft AI for Good Research Lab; School of Public Health, Brown University; Harvard T.H. Chan School of Public Health; Brigham and Women’s Hospital

## Abstract

**Importance:** With an abundant supply of COVID-19 vaccines becoming available in spring and summer 2021, the major barrier to high vaccination rates in the United States has been a lack of vaccine demand. This has contributed to a higher rate of deaths from SARS-CoV-2 infections amongst unvaccinated individuals as compared to vaccinated individuals. It is important to understand how low vaccination rates directly impact deaths resulting from SARS-CoV-2 infections in unvaccinated populations across the United States.

**Objective:** To estimate a lower bound on the number of vaccine-preventable deaths from SARS-CoV-2 infections under various scenarios of vaccine completion, for every state of the United States.

**Design, Setting, and Participants:** This counterfactual simulation study varies the rates of complete vaccination coverage under the scenarios of 100%, 90% and 85% coverage of the adult (18+) population of the United States. For each scenario, we use U.S. state-level demographic information in conjunction with county-level vaccination statistics to compute a lower bound on the number of vaccine-preventable deaths for each state.

**Exposures:** COVID-19 vaccines, SARS-CoV-2 infection

**Main Outcomes and Measures:** Death from SARS-CoV-2 infection

**Results:** Between January 1st, 2021 and April 30th, 2022, there were 641,305 deaths due to COVID-19 in the United States. Assuming each state continued peak vaccination capacity after initially achieving its peak vaccination rate, a vaccination rate of 100% would have led to 322,324 deaths nationally, that of 90% would have led to 415,878 deaths, and that of 85% would have led to 463,305 deaths. As a comparison, using the state with the highest peak vaccination rate (per million population each week) for all the states, a vaccination rate of 100% would have led to 302,344 deaths nationally, that of 90% would have led to 398,289 deaths, and that of 85% would have led to 446,449 deaths.

**Conclusions and Relevance:** Once COVID-19 vaccine supplies peaked across the United States, if there had been 100% COVID-19 vaccination coverage of the over 18+ population, a conservative estimate of 318,981 deaths could have been potentially avoided through vaccination. For a 90% vaccination coverage, we estimate at least 225,427 deaths averted through vaccination, and at least 178,000 lives saved through vaccination for an 85% vaccination coverage.

## 1 Introduction

In early 2021, a little over one year after the emergence of the novel virus SARS-CoV-2, effective vaccines became available in the United States. Historic efforts in vaccine manufacturing and distribution enabled rapidly growing supplies. By mid-April 2021, vaccine availability outstripped demand and daily vaccination rates peaked across the nation. By summer 2021, U.S. vaccination rates lagged behind many Western nations [1].

Clinical trials as well as large-scale observational data in the U.S. demonstrate that vaccines are highly effective in preventing severe disease, hospitalization, and death from COVID-19 infections [2] [3].

Of the 641,305 deaths recorded in the U.S. between January 1st 2021 and April 30th 2022, only 59,518 were in fully vaccinated individuals, while 581,786 were not [4]. In other words, 90.7 percentage of deaths in this timeframe occurred in unvaccinated or only partially vaccinated Americans.

In Figure 1, we show a visual comparison of the reported death rate between fully vaccinated vs. unvaccinated/incomplete course adult (18+) populations in the U.S. during 2021, illustrating that the death rate is significantly higher amongst unvaccinated populations.

**Figure 1:**
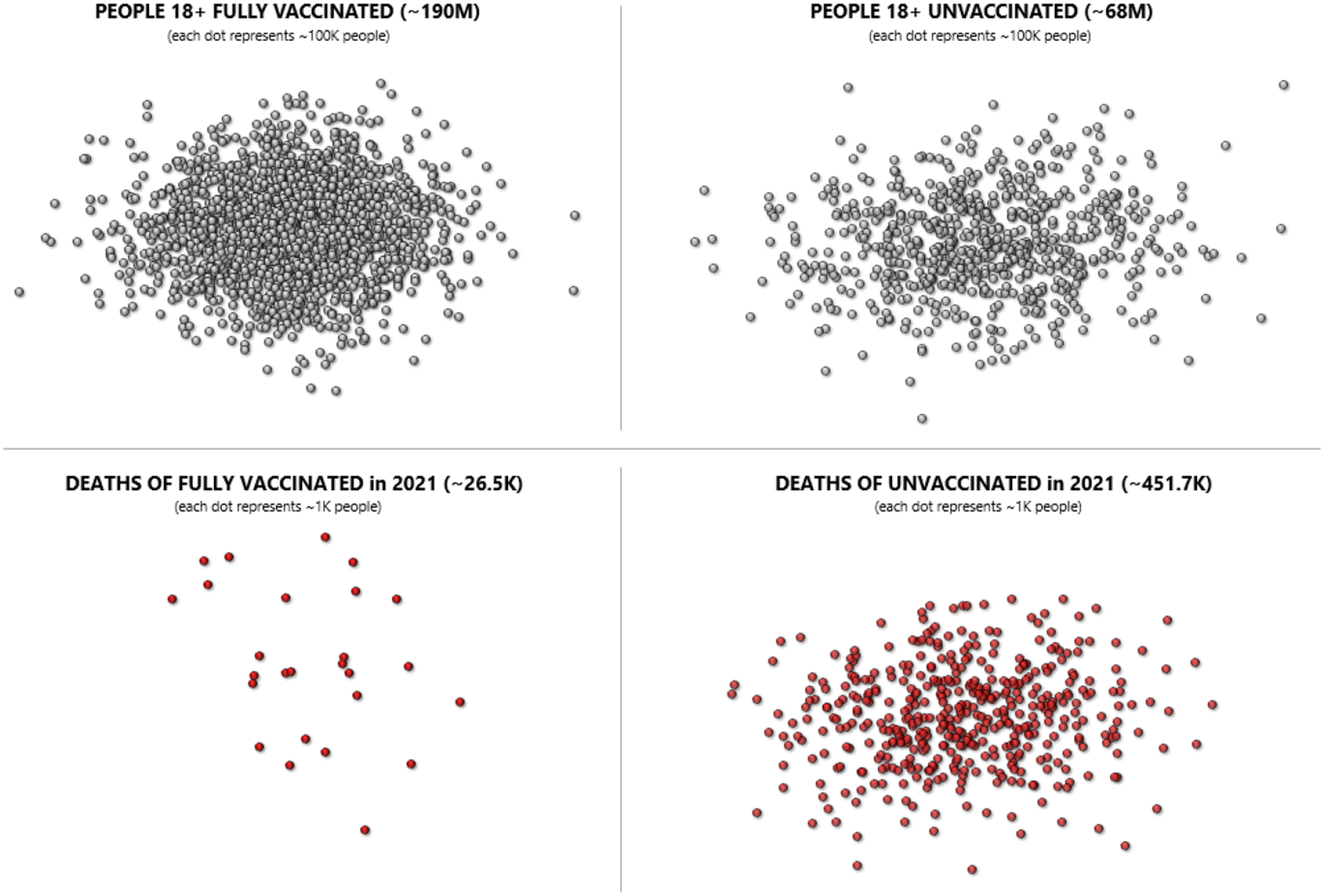
This figure shows a comparison of the magnitudes of vaccinations and deaths among vaccinated and unvaccinated populations. Each grey dot represents 100,000 people, each red dot represents 1,000 people. The placement of the dots in each panel is for illustration only.

Given the demonstrated efficacy of vaccines, it is instructive to estimate how many lives might have been saved by higher vaccination rates in the U.S. adult population. This can help policy makers better understand the cost of incomplete vaccine coverage and inform appropriate prioritization of resources in future vaccination efforts against COVID and other diseases.

To explore answers to this question, we built a counterfactual or “what if” model that simulates various rates of two-dose vaccination and their impact on death rates. We project completed vaccinations if demand for vaccines had remained at its peak, and been met consistently by supply until target rates of vaccination were achieved.

Our counterfactual scenario estimates the number of deaths in individuals who were not fully vaccinated in the period after an adequate supply of vaccines became available in their state, based on the number of recorded Covid-19 deaths that occurred between January 1, 2021 and April 30, 2022. Note that with limited and incomplete information, our methodology estimates a reasonable and reliable lower bound on deaths that may have been preventable had vaccination rates in the U.S. been higher. Our estimates should be viewed as a conservative benchmark.

Based on the assumptions described in the Methods section, we estimate the counterfactual death number for each state in three distinct scenarios: by assuming either 85%, 90% or 100% of adults to be fully vaccinated in each given state. Estimating vaccine-preventable deaths in different scenarios and for different geographic areas allows a closer look at how lack of vaccine demand is impacting Covid-19 mortality, and can benefit local, state and federal government in their decision making about vaccination policies, programs and funding.

By April 30, 2022, approximately 24 percent of adults and 57 percent of children in the U.S. remained unvaccinated, leaving 30 percent, or around 92 million people vulnerable to severe complications and death from Covid-19.

Prior work estimating the number of lives lost to under-vaccination has been limited in scope by obtaining coarse, nationwide aggregate estimates [5] [6] or state-level estimates for specific states [7], for a fixed scenario of vaccine utilization [8], being restricted to select racial groups [9], or has relied on the number of excess deaths as the input data. Other “what if” analyses condition on nationally uniform vaccine uptake, without considering state-level supply constraints [6]. Our work is based on a counterfactual model that conditions on vaccine availability at the state level, uses comprehensive regional data from CDC, and derives number of deaths due to under-vaccination under three different scenarios for every state in the U.S.

## 2 Methods

### 2.1 Study population

We consider the population of the US that is captured by the following three datasets that report COVID-19 cases, deaths, and vaccinations at various regional levels.

1. “Coronavirus (Covid-19) Data in the United States - Cumulative Cases and Deaths” released by New York Times (2022): this dataset includes state-level vaccination completion and death count, and is updated daily [10].
2. “COVID-19 Vaccinations in the United States” released by CDC (2022): this dataset includes the number of adults (age 18+) who have been fully vaccinated for each state. We also use this data to infer the adult (age 18+) population for each state [4]. The data is updated daily.
3. “Rates of COVID-19 Cases and Deaths by Vaccination Status” released by CDC (2022): this dataset includes death rates of COVID-19 by vaccination status (that is, fully vaccinated vs. unvaccinated or partially vaccinated) [11]. The data starts from April 2021 and is updated weekly. It is collected at a national level, so we will assume the same death rate for each state.

### 2.2 Statistical methods

With the datasets described above, our approach is based on the following assumptions:

1. We assume that if a state had the capacity to vaccinate X number of people per day at one point in the pandemic, that state could have maintained this peak capacity given enough demand. Indeed, given enough demand, states would likely have continued to increase their vaccination capacity. This assumption therefore provides an underestimate of vaccination capacity and thus an underestimate of the counterfactual preventable deaths (See Figure 2). Since most states had the ability to increase capacity beyond their demand-driven peak vaccination rate, we also estimated the counterfactual deaths for a second scenario, in which we apply the rate of vaccination in the state with the highest peak vaccination rate (per million population each week) to all other states who did not attain this peak, but potentially might have given sufficient demand. This scenario offers insights on the impact of a faster vaccination pace beyond an individual state’s peak. (The state with the highest vaccination rate in the nation was Maine during the week of April 11, 2021.)

**Figure 2:**
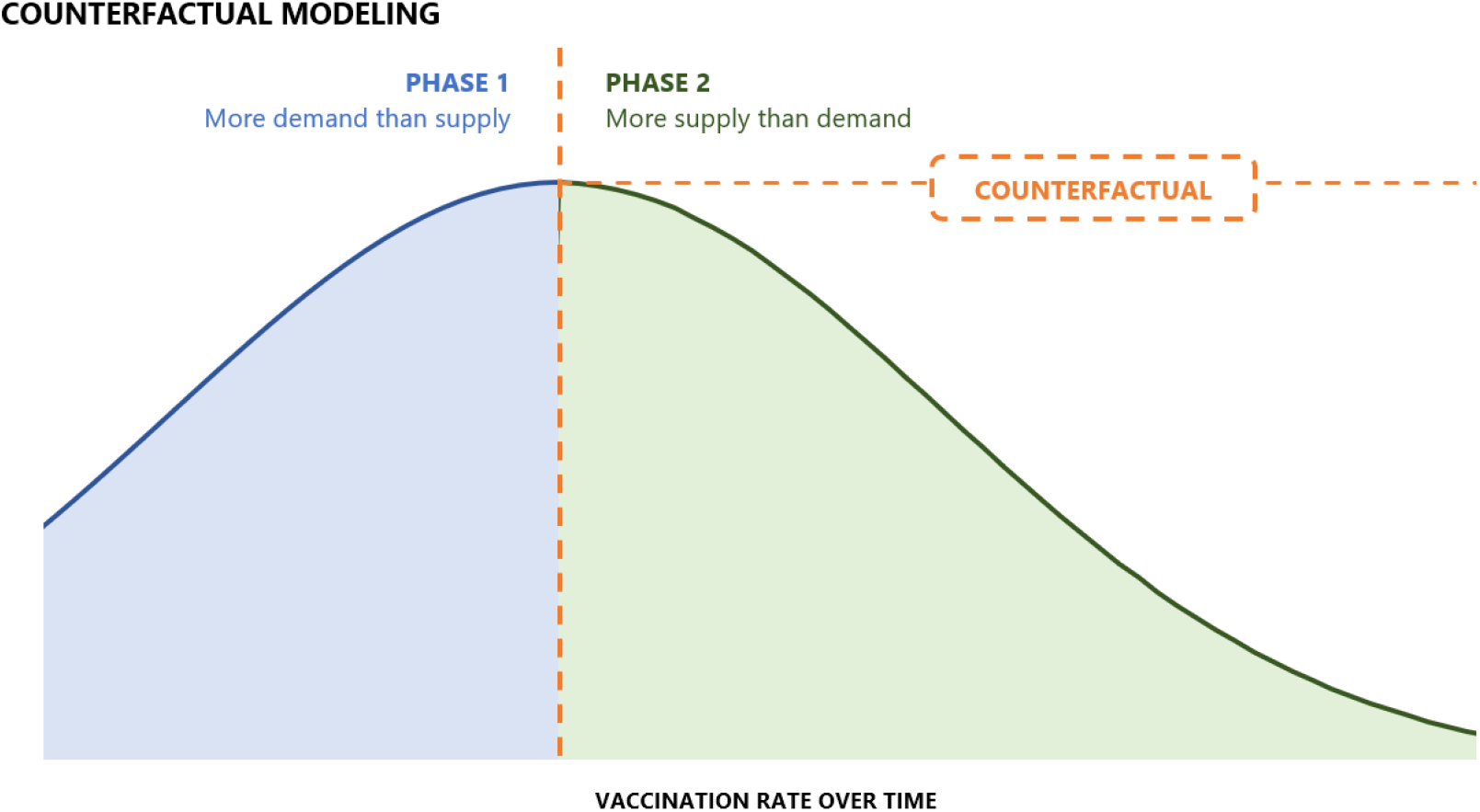
Counterfactual modeling for preventable deaths due to COVID-19 during the phase when there was more supply than demand.
2. We assume no change in the effective reproductive number of COVID-19 cases. As more people get vaccines, the effective reproductive number should become smaller due to the positive externality of vaccinations, but we do not take this factor into account. This assumption will result in underestimating counterfactual preventable deaths. As people’s behavior may change over time (for example, more outdoor activities and travels during summer times and holidays), the effective reproductive number is expected to fluctuate.
3. Although the CDC publishes vaccine effectiveness by age, we do not have vaccine completion breakdowns by age in our data. Therefore, to keep the level of information consistent across sources, we used the weekly average of vaccine effectiveness reported for all individuals aged 18+ (see Figure 3). This leads to the assumption that the vaccine effectiveness is the same for all individuals regardless of their age.

**Figure 3:**
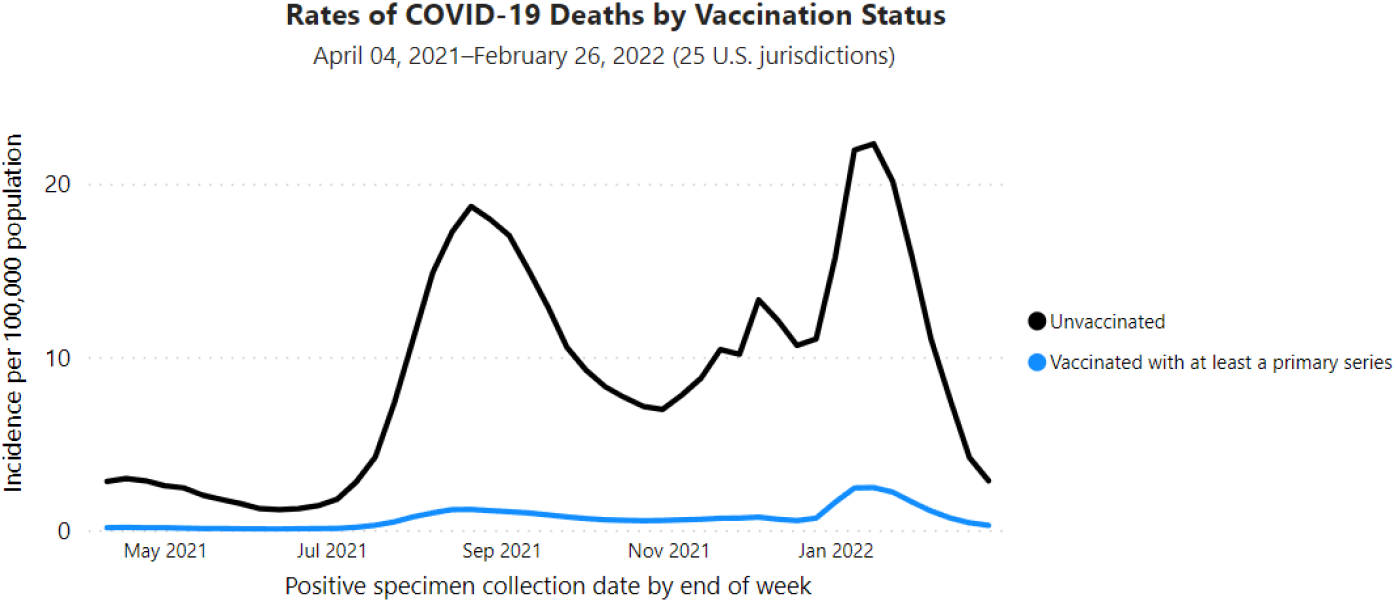
Weekly average death incidence (per 100,000 population) comparison between unvaccinated group vs. fully vaccinated group, for all ages, all vaccination products, and all states. The chart is released by CDC and is available at https://covid.cdc.gov/covid-data-tracker/#rates-by-vaccine-status
4. Since we do not have detailed data for all age-cohorts at the county level, we assume that the age distribution of vaccinated and unvaccinated groups remains the same in our counterfactual scenario, and every unvaccinated individual has an equal likelihood of getting vaccinated. We know that the first group to get prioritized to be vaccinated were the older cohorts, who are also the ones with the highest death rate. In the counterfactual scenario in which vaccine demand is maintained, those cohorts would have been vaccinated first-likely averting more actual deaths. But our model assumes a constant age mix resulting in an underestimation of counterfactual preventable deaths (see Table 1).

**Table 1:** Notations used in the formulation of estimating counterfactual deaths.

| NOTATION | MEANING |
| --- | --- |
| $t$ | With $t = 0$ indicating the week when the peak of vaccination was achieved. |
| $D_{\{e,a\},t}$ | Number of deaths at week $t$; $e$ = estimated, $a$ = actual. |
| $R_{\{v,u\},t}$ | Death rate at week $t$; $v$ = vaccinated, $u$ = unvaccinated. |
| $P_{18+}$ | Population aged 18 and above, for a given state. |
| $V_0$ | Number of vaccines administered at the peak (that is, at week $t = 0$). |
| $V_t$ | Estimated total number of vaccinated people as of week $t$. |
| $U_t$ | Estimated total number of unvaccinated people as of week $t$. |
| $c$ | target percent (100%, 90%, or 85%) of total population that is vaccinated. |

The above assumptions simplify the complex estimation problem and let us obtain a lower bound estimate of the counterfactual deaths. Table 1 contains the notation used in our computation. The logic behind our counterfactual estimation method is straightforward and can be illustrated as follows: For a given state *S*, let the week when the vaccine administration for this state reached its peak be encoded as time point *t* = 0. This marks the starting point of the counterfactual simulation. Thus, until the week *t* = 0, the estimated number of weekly deaths *D*(*e, t*) is the same as the actual number of weekly deaths *D*(*a, t*). Mathematically, we can write:

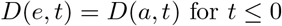

Given the number of vaccines *V*_0_, administered at the peak (i.e., at week *t* = 0), we can establish the following recurrence to get the subsequent estimates (i.e., the estimates for weeks *t >* 0) for the vaccinated population: *V* (*t*) and the unvaccinated population: *U* (*t*) for every state:

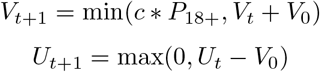

We compute the above recurrence until the estimated total number of fully vaccinated adults reaches the target percentage c of population in each state. The weeks starting from *t* = 1 until the week where the target vaccination percentage is reached are considered as the “counterfactual window” for the given state.

Next, for the weeks *t >* 0, following the “peak” week, given the death rate *R*_*v,t*_ of the fully vaccinated cohort vs. the death rate *R*_*u,t*_ of the unvaccinated cohort at week *t*, we can calculate the estimated weekly death count for each state using the following equation.

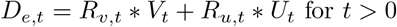

Finally, the total preventable deaths for a given state are obtained by aggregating the difference in the estimated deaths and actual deaths over all the weeks in the counterfactual window. This is given by,

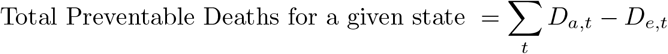

We repeat this entire computation for all states and obtain the corresponding total estimates of preventable deaths in the United States.

## 3 Results

Using the approach described above, we simulated outcomes for three scenarios of vaccine completion rates and estimated the counterfactual deaths under each scenario for the time between January 1st, 2021 and April 30th, 2022 (see Table 2).

**Table 2:** Counterfactual death estimation results for the assumptions of 100%, 90% and 85% vaccination completion, respectively.

| ASSUMING EACH STATE CONTINUED ITS MAXIMUM VACCINATION CAPACITY<br>AFTER ACHIEVING THE PEAK VACCINATION RATE |  |  |  |
| --- | --- | --- | --- |
| ASSUMED PERCENTAGE<br>OF FULL VACCINATION | NUMBER OF NON-VACCINE-PREVENTABLE DEATHS<br>ESTIMATION | NUMBER OF VACCINE-PREVENTABLE DEATHS<br>ESTIMATION | PERCENTAGE OF VACCINE<br>PREVENTABLE DEATHS |
| 100% | 322,324 | 318,981 | 50% |
| 90% | 415,878 | 225,427 | 35% |
| 85% | 463,305 | 178,000 | 28% |
| ASSUMING ALL THE STATES HAD REACHED THE SAME VACCINATION RATE<br>ACHIEVED BY THE STATE WITH THE HIGHEST RECORDED PEAK VACCINATION RATE (MAINE; APRIL 11, 2021) |  |  |  |
| ASSUMED PERCENTAGE<br>OF FULL VACCINATION | NUMBER OF NON-VACCINE-PREVENTABLE DEATHS<br>ESTIMATION | NUMBER OF VACCINE-PREVENTABLE DEATHS<br>ESTIMATION | PERCENTAGE OF VACCINE<br>PREVENTABLE DEATHS |
| 100% | 302,344 | 338,961 | 53% |
| 90% | 398,289 | 243,016 | 38% |
| 85% | 446,449 | 194,856 | 30% |

On average, 50% of deaths were preventable assuming 100% full vaccination at weekly maximum rate. However, there is quite a large variance among states – ranging from 25% to 74%. The actual vaccination percentage and when the peak week occurred contribute to this variance. For example, Washington D.C., Massachusetts, Puerto Rico and Vermont stand out as having the smallest percentages of avoidable deaths thanks to robust vaccine rollout. In contrast, West Virginia, Wyoming, Tennessee and Kentucky are among the highest percentages of avoidable deaths.

In examining two of the largest states by population, we compared California and Florida to assess how their vaccination trajectory related to vaccine-preventable deaths. See Figure 4 for the avoidable deaths comparison between these two states over time assuming 100% vaccination. Figure 5 shows vaccine preventable deaths per million (18+) assuming 100% vaccination rate for all states.

**Figure 4:**
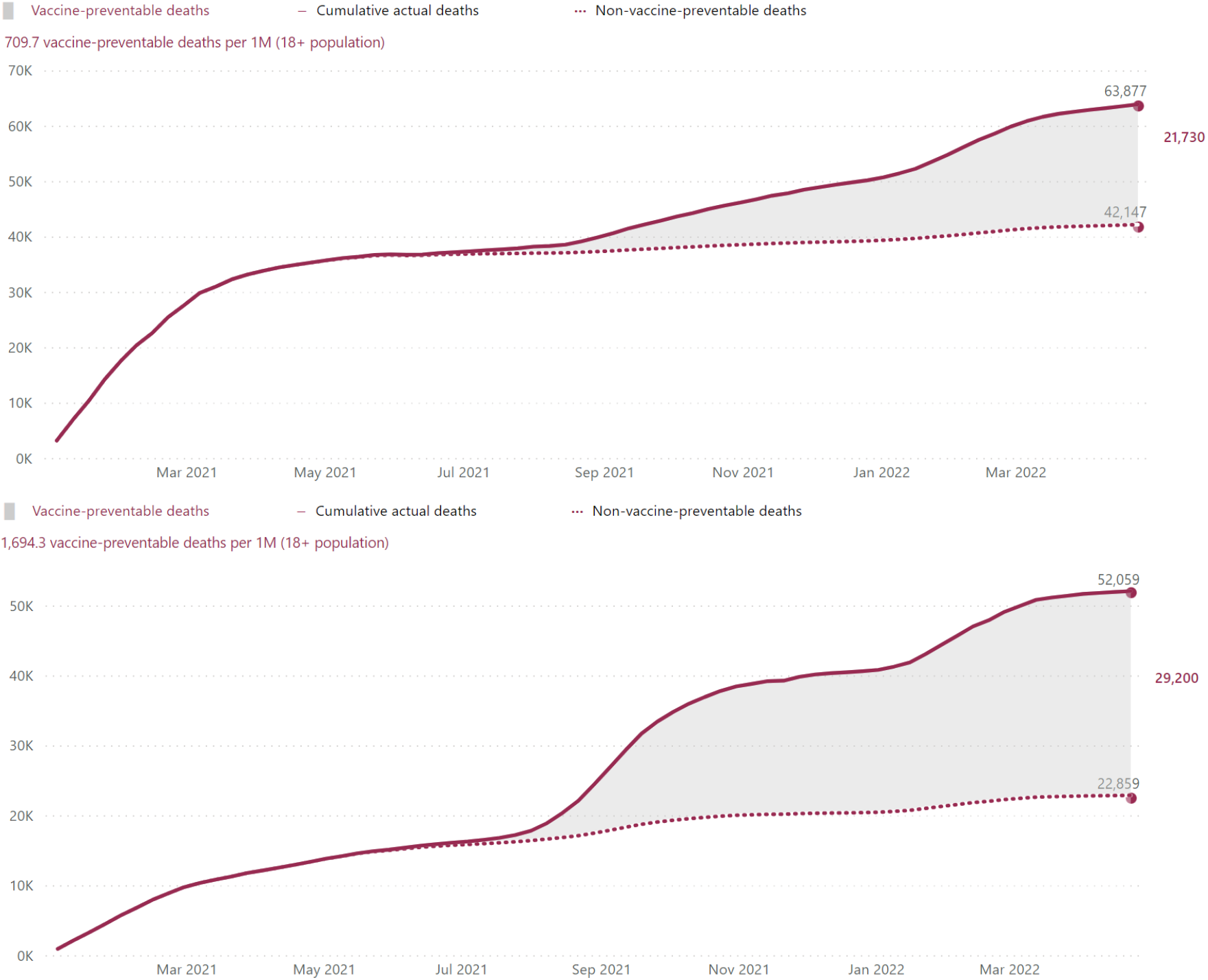
Percentage of preventable deaths in the United States between January 1st, 2021 and April 30th, 2022 by assuming 100% vaccine completion for California (top) and Florida (bottom), respectively.

**Figure 5:**
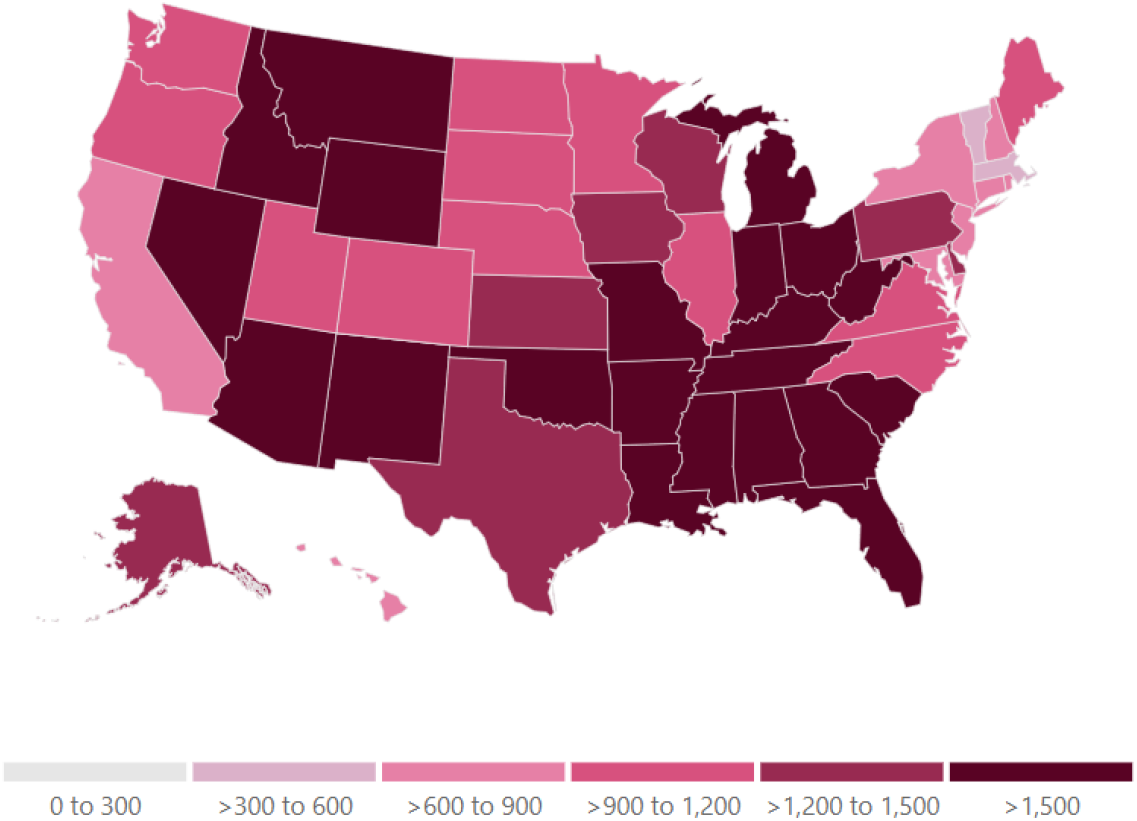
Preventable deaths per million (18+) in the United States between January 1st, 2021 and April 30th, 2022 by assuming 100% vaccine completion at maximum vaccination capacity.

These results partially reflect how each state made their own decisions on balancing its social, public health and economic priorities, as well as how much to invest in vaccine access and demand. While the discussion of policy making goes beyond the scope of this paper, our estimates are but one input policy makers may consider in future decisions.

## 4 Conclusion

In a national analysis of three counterfactual scenarios of higher-than-observed full vaccination rate, we find that that increased vaccination in the U.S. could have potentially averted approximately 178,000 to 318,981 deaths between January 1st, 2021 and April 30th, 2022. Our analysis adds to the growing literature studying how lack of vaccine demand impact individuals, health systems, and the society at large. These estimates provide an empirical estimate of the public health costs associated with suboptimal vaccination coverage of the population and may guide policymakers in optimizing vaccination rates in states where vaccine preventable deaths numbers are high.

## Data Availability

All data produced in the present study are available upon reasonable request to the authors.

https://data.cdc.gov/Public-Health-Surveillance/Rates-of-COVID-19-Cases-or-Deaths-by-Age-Group-and/3rge-nu2a/data

https://github.com/nytimes/covid-19-data

